# Exploring the Temporal Dynamics of County-Level Vulnerability Factors on COVID-19 Outcomes

**DOI:** 10.1101/2021.11.24.21266757

**Authors:** Jing Zhang, Daesung Choi, Shivani A Patel, Joyce C. Ho

## Abstract

As the outbreak of COVID-19 has become a severe worldwide pandemic, every country fights against the spread of this deadly disease with incredible efforts. There are numerous researches along with every conceivable dimension for COVID-19. Among these researches, different demographic and contextual factors of populations and communities also play an essential role in providing more information for decision-makers. This paper mainly utilizes existing data on county contextual factors at the United States county-level to develop a model that can capture the dynamic trajectory of COVID-19 (i.e., cases) and its impacts across the United States. Moreover, our methods applied to contextual data achieves better results compared with existing measures of vulnerability.

## 1 Introduction

Since December 2019, there have been 252 million confirmed cases of the coronavirus disease-19 (COVID-19) [3], with total deaths reaching five million. COVID-19 brought an unprecedented social and economic impact worldwide, and it is still accelerating without presenting any signs of nearing an end. A lot of prodigious COVID-19 projects have been conducted by researchers in fields ranging from epidemiology to economics. At the same time, many states in the United States imposed policies to control the spread [22]. There are several projects focused on forecasting and modeling the disease in near real-time [5, 17]. Yet identifying and selecting the proper factors to construct community vulnerability indices that can be used to understand and mitigate disease spread remains relatively understudied [7, 15]. As a motivating example, different communities in the United States are susceptible to varying severities of adverse effects when faced with a public health crisis [19]. Identifying the vulnerable areas can help the public health department utilize proper measures to mitigate these negative impacts.

The Centers for Disease Control and Prevention (CDC) provided the information about the level for the COVID-19 travel health notices [2]. It is also an indicator about the COVID-19 status of the destination, and according to the criteria, people will obtain a better sense about the risk of COVID-19 infection. It utilized the incidence rate and case count thresholds to assign the level of the notice. For example, the incidence rate over past 28 days is applied for the destinations with populations above 100,000; otherwise, the case count is used. If the rate or count of the destination is more than 500, the place will be in level 4 (very high); if it falls into the range between 100 and 500, the place will be in level 3 (high); if it is less than 100 but higher than 50, the place will be in level 2 (moderate), and if it is less than 50, the place will be noticed as level 1 (low).

The CDC also tried to leverage the social vulnerability index (SVI) to identify the vulnerable areas. SVI ranks census tracks using four themes: socio-economic status, household composition, minority status and language, and access to housing and transportation [8]. Unfortunately, SVI does not provide comprehensive determinations of vulnerability for the COVID-19 situation since it was designed primarily for natural disaster crises [11]. Similarly, the Robert Wood Johnson Foundation (RWJF) produced the County Health Rankings with 19 factors from census and American Community Survey [18]. Yet these rankings are also not suitable for identifying vulnerable areas in the COVID-19 pandemic.

Recently, the CDC and Surgo Foundation proposed the COVID-19 Community Vulnerability Index (CCVI) [9] that uses a statistical linear algorithm on 29 factors, which failed to capture the nonlinearity of the vulnerability. In [21], the public health planners and policymakers was interested in a more complex methodology to capture the vulnerability along with the dynamic trajectory of COVID-19. Therefore, they developed a new vulnerability index - C19VI, which quantifies each county’s vulnerability with the same features from CCVI. However, instead of using a linear statistical algorithm, they implemented Random Forest [20] to calculate the C19VI using two stages. First, it applied homogeneity analysis [16] and trend analysis [14] to obtain the COVID-19 impact score and rank, which were used to annotate the counties with different levels. Next, based on the annotation for the counties, it generated the C19VI by Random Forest. Compared with CCVI, C19VI introduced a new way to utilize the infection fatality rate, cases and deaths. However, there are two major limitations. The definition of the six impact groups is inconsistent with that from the CDC Travel Health Notice [2], it leans more on mortality. The result of C19VI on October 31, 2020 shows that 83% of the counties are in the low impact status, however, 79% of the counties are above the high-level status based on CDC travel health notices. If we utilize the result of C19VI, it will potentially expose people to the risks.

Li et al. [12] utilized 23 county-level features related to social demographics, population activities, mobility, social network structure, and disease-related attributes and the COVID-19 pandemic data from March 24 to June 23, 2020, to capture the dynamic impacts within these features. While the authors provide an excellent visualization illustrating the dynamic impact of the features along with the trajectory of the pandemic, it also suffers from two weaknesses. (1) it grouped the counties into five classes, where the first class contained all counties with zero new cases, and the rest of the counties were evenly split by the daily new cases. This group strategy is unrealistic as grouping based on daily cases will not yield a uniform distribution. (2) the predictive results are poor (∼ 46%), calling into question the generalizability of the ranked features.

We propose to incorporate new county-level factors in concert with the CCVI features to yield more informative models that can better guide public health planners and policymakers. Moreover, we train XGBoost models [6] based on the CDC Travel Health Notice levels on a monthly basis to (1) better understand the temporal dynamics of county-level features on COVID-19 outcomes and (2) to anchor the feature importance with public health guidance. Our results demonstrate that the new feature set outperforms existing vulnerability index features by a significant margin. Due to the page limitation, we only present the feature importance along with the modified trajectory of COVID-19 rather than creating a new index, but note that our analysis leads to an improved vulnerability index.

## 2 Approach

We utilize the publicly available processed datasets from Emory University [1] to obtain the monthly data for 3142 counties in the United States from April 2020 to August 2021. We obtained the ‘static’ contextual and CCVI features from Surgo Foundation [9], United States Department of Homeland Security [10], and the United States Census Bureau [4]. This data was used to extract the 19 RWJF features, the 29 CCVI features, and two additional features (*traffic* and *Air Pollution*). This yields a final feature set of 41 factors. We excluded the 36 counties with missing values (i.e., final number of counties is 3106).

Each datapoint for each county is assigned a label based on the CDC travel notice level. 12 counties were tagged with a very high level travel notice 14 times in the past 17 months. We also note that the numbers of different class samples vary across the different months. For example, in August 2020, there were 1076 counties in low level, 478 counties in moderate, 1223 counties in high level, and 329 counties in very high level. One year later (August 2021), there were 397, 234, 1108, and 1367 counties labeled low, moderate, high, and very high respectively.

We utilized XGBoost rather than Random Forest to return a concise version of feature importance by accounting for correlated features. Due to class imbalance, the micro F1 score was utilized for the evaluation metric. 623 counties are used as the test set with the remainder serving as the training set. Since most of the features are static, we created a static baseline model where XGBoost was trained with 17 months of data for 2483 counties and then evaluated on the 17 months of data for the 623 counties. The remaining experiments involved only the monthly data. For example, we utilized the 2483 counties data in August 2020 to train a model and test it on the 623 counties data in August 2020. We then identify the most critical features for that month using the feature importance. For all the experiments, we benchmarked our feature set with RWJF and CCVI. For each experiment, we will run five times to obtain the stability of the model.

## 3 Results

The first two rows in Table 1 summarizes the mean F1 score across the 17 months for the different feature sets and modeling assumptions. We note that the static baseline, which assumed feature importance was fixed across the pandemic, yielded a high F1 score of 0.4942. In comparison, modeling feature importance on a monthly basis yielded a ∼ 45 − 60% performance improvement across the three feature sets. The significant performance increase demonstrates the importance of modeling the temporal dynamics of vulnerability factors.

**Table 1:**
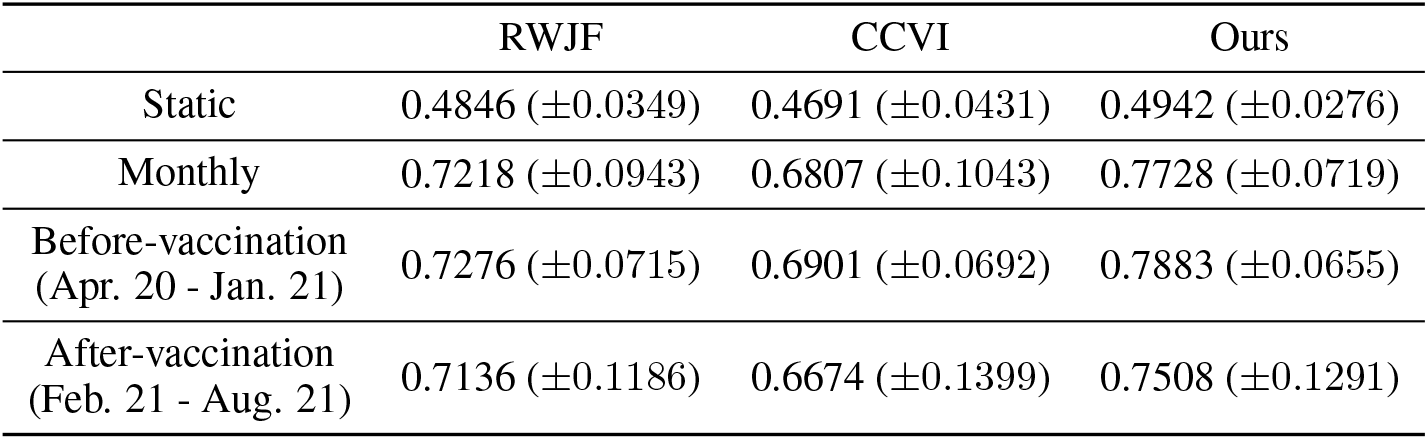
Results for the different feature sets (Mean F1 score with standard deviation)

|  | RWJF | CCVI | Ours |
| --- | --- | --- | --- |
| Static | 0.4846 ($\pm 0.0349$) | 0.4691 ($\pm 0.0431$) | 0.4942 ($\pm 0.0276$) |
| Monthly | 0.7218 ($\pm 0.0943$) | 0.6807 ($\pm 0.1043$) | 0.7728 ($\pm 0.0719$) |
| Before-vaccination<br>(Apr. 20 - Jan. 21) | 0.7276 ($\pm 0.0715$) | 0.6901 ($\pm 0.0692$) | 0.7883 ($\pm 0.0655$) |
| After-vaccination<br>(Feb. 21 - Aug. 21) | 0.7136 ($\pm 0.1186$) | 0.6674 ($\pm 0.1399$) | 0.7508 ($\pm 0.1291$) |

Table 1 also illustrates that our feature set offers better predictive power than RWJF and CCVI. Regardless of the static or monthly modeling, there is a performance boost compared to the other two feature sets. The 19 RWJF factors provide considerable boost over the 29 CCVI features. When we remove the two added features (Air Pollution and Traffic) from our feature set, the monthly F1 score drops to 0.7519. We also assessed the feature sets by dividing the pandemic into two stages, pre-vaccination and post-vaccination, which is shown in the last two rows of Table 1. The factors achieved slightly higher performance across all three feature sets for pre-vaccination compared to post-vaccination. Moreover, even though we did not include any features related to vaccination, our feature set doesn’t have a significant decrease in performance between the two stages. From the results of the standard deviation for five-time experiments, we can see that our feature set could provide more stable results comparing with other feature sets.

Figure 1 illustrates the feature importance for each month. Factors like ICU beds, Uninsured, Traffic, Air Pollution, and Urbanization dominate throughout the pandemic. We observe several features including Age over 65, Minority, and Mobile Homes, exhibit several behavior changes during the pandemic, suggesting that these features have different temporal impacts. Moreover, Mobile Homes has a periodic behavior where its importance peeks during the summer months. To be more specific, we examined the feature of ICU beds in 2019. There are 2290 counties which own less than ten ICU beds, 61% of the counties are in the status above high level. 92% of the counties which have more than 10 ICU beds are in the status above high level. This observation explains why ICU beds plays an important role in the classification task.

**Figure 1:**
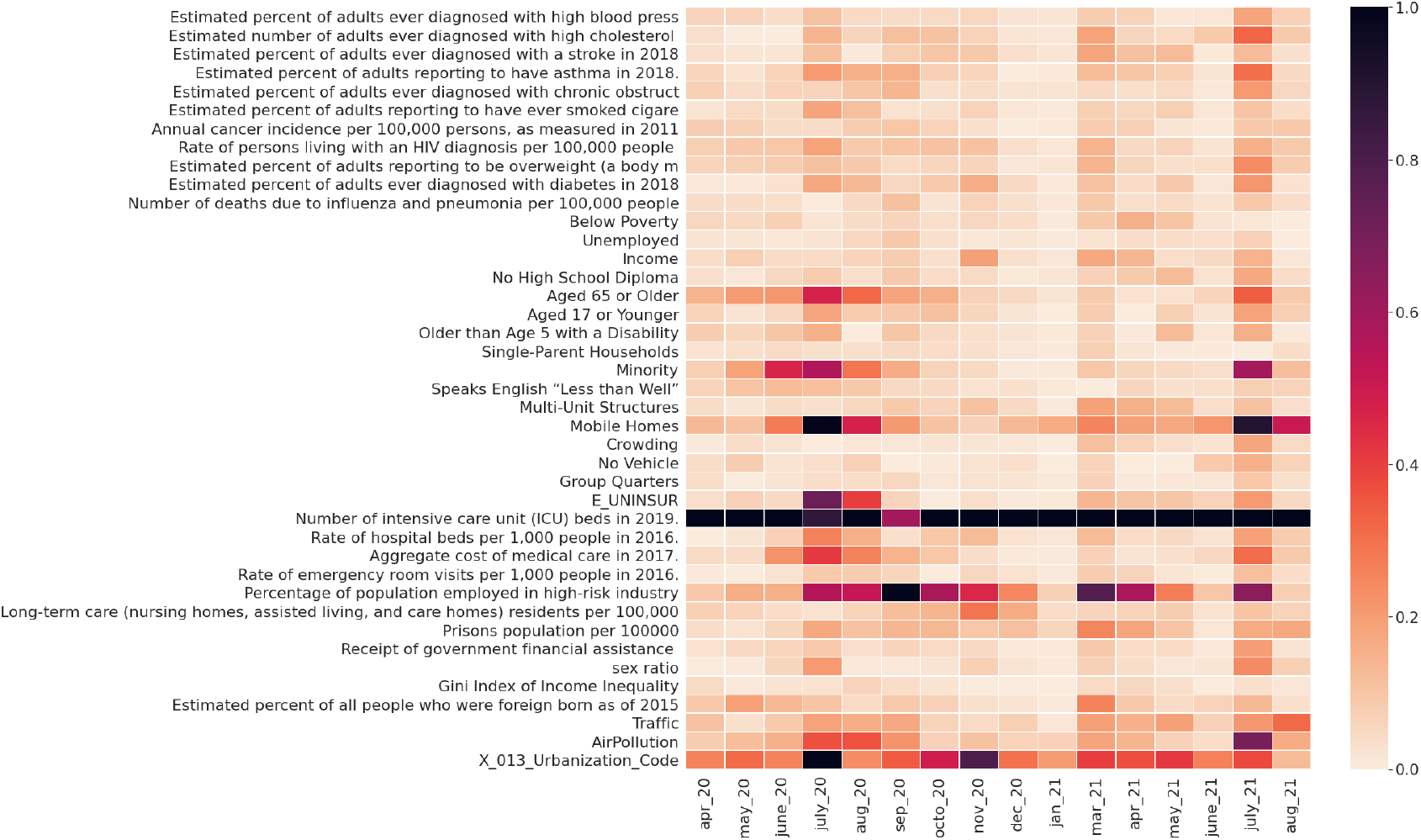
Feature importance along with months

We also leveraged SHAP [13] to visualize the contributions of each feature to different classes. As shown in Figure 2, we can see that even though the feature of ICU beds dominates in May 2020 and 2021, it has different impacts on different classes. For example, in May 2020, the ICU beds factor affects the High-Level class more than others, whereas in May 2021, the impact is on Low-Level class. This illustrates the importance of understanding the temporal dynamics associated with each of the travel notice classes. Moreover, from Figures 1 and 2, we observe the important role the two new features (Air Pollution and Traffic) play in the prediction.

**Figure 2:**
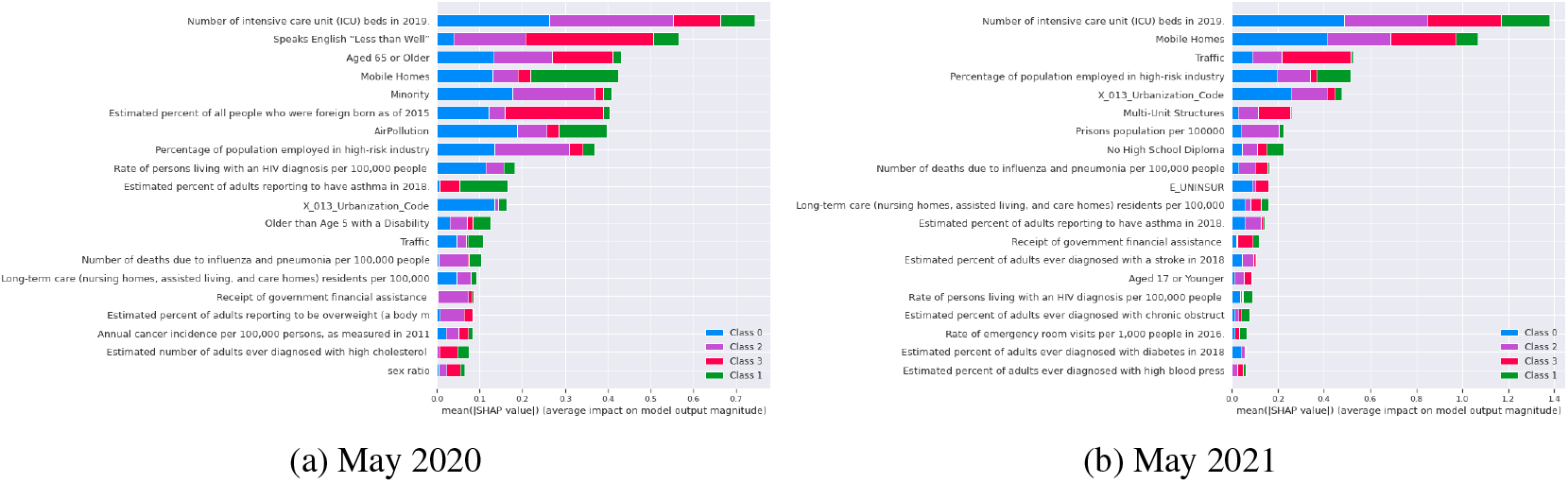
Feature contributions to the different travel notice classes, where 0 (blue), 1 (purple), 2 (red) and 3 (green) correspond to low, moderate, and very high level respectively.

## 4 Conclusion

This paper leveraged the XGBoost model with 41 features to conduct a four-class monthly classification task and obtained an improved F1 score compared to other feature sets. By allowing the dynamics to change on a monthly basis, our features can better capture the trajectory of COVID-19.

The model provides evidence that vulnerability is dynamic and therefore opens the door for new approaches. This enables the decision-makers to formulate a coherent response to mitigate the negative impacts for vulnerable areas. We plan to extend our promising exploration results to build a more comprehensive COVID-19 vulnerable index. On the other hand, automating the data acquisition and analysis for dynamic analysis could be useful for policy makers so that their decisions are based on best available data. That is, this is encouraging a paradigm shift in how we incorporate social vulnerability into estimation.

## Data Availability

All data produced are available online at COVID 19 Emory dashboard

https://covid19.emory.edu

## Acknowledgements

This work was supported by the National Science Foundation award IIS-#1838200, National Institute of Health award 1K01LM012924, Robert Wood Johnson Foundation #77624, Emory Covid-19 Response Collaborative, and Google Cloud Platform research credits.

